# Ancient DNA confirms anaemia as the cause for Porotic Hyperostosis in ancient Neolithics together with a genetic architecture for low bone mineral density

**DOI:** 10.1101/2023.01.11.23284324

**Authors:** Manuel Ferrando-Bernal

## Abstract

Porotic hyperostosis is a disease that used to have an important prevalence during the Neolithic. Several hypotheses have been described to explain its origin but not one has been tested genetically. Here, I used hundreds of SNPs to confirm anaemia and low bone mineral density as the main cause for this disease using data for 80 ancient individuals for which it is known if they had or not the disease. Additionally, Neolithic individuals show the lowest bone mineral density and haemoglobin levels of all other periods tested here, explaining the highest prevalence of the porotic hyperostosis during this age.

## 1. Introduction

Paleopathology is the study of disease in ancient organisms. Most of its studies focused on past pathologies affecting ancient human remains (1,2). Due to the intrinsic nature of ancient remains bone diseases are the best well studied in paleopathology such as bone cancer, rickets or bone traumatisms. One of the most known diseases affecting past populations is Porotic hyperostosis (PH) which is a skeletal disorder characterised by bony lesions on the bones of the skull due to an expansion of the cranial diploë (3) (Figure 1). This condition has been observed in all ancient populations (4-8), but mainly during the Neolithic period (2, 9-11), being observed in 43% of the individuals in some populations (11). The lesions are typically found on the frontal and parietal bones, and can range from small pits to large lesions. The main cause behind this disease is yet not well known, likely due to its low prevalence in modern times (12,13). Many hypotheses have been suggested. For example, low vitamin D levels may lead to thinner skulls due to its implication in skeletal mineralization (6). Another hypothesis is nutritional deficiency at weaning during childhood (4). However, since the 1950s, anaemia has been thought as the main cause for PH (14). It has been linked to infectious disease, such as malaria, as the parasite *Plasmodium falciparum* might have caused anaemia by an excess of red blood cell destruction (15). Also, low vitamin B12 has been proposed, as its deficiency may cause red blood cells abnormalities and then a need for a diploë expansion to increase the red cell production (14). Also anaemia caused by thalassemia has been suggested as the main cause for PH (13, 15) and even an iron-deficiency anaemia, caused by a lack of dietary iron, poor absorption of iron, or increased demand for iron due to high levels of physical activity (14). Additionally, Eurasians seem to have been selected for low bone mineral density levels during the last thousand years (16,17), which may lead to particular thin bones in some individuals.

**Figure 1.**
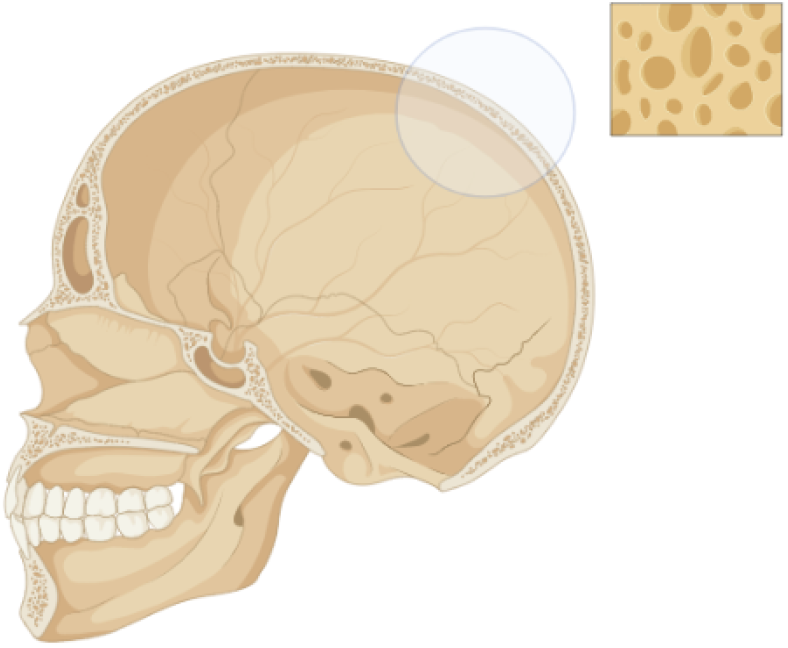
Porotic hyperostosis is characterised by ticker skull bones with an increased porous area caused by an expansion of the cranial diploë.

To date, studies of PH in ancient individuals used indirect evidence to test for a particular hypothesis or another. However, present-day technologies and more than three decades of study of ancient DNA allow us to test them directly. To date, we have been able to recover the genomes of hundreds of ancient samples and test many hypotheses that have been unanswered for decades, such as our past relationships with other *Homo* species (18, 19), connections among ancient and modern individuals (20) or the selective pressures of particular human traits (21). Despite some studies focused on how breeding with Neanderthals and Denisovans affect the development of different diseases in modern individuals (22), the contribution of Ancient DNA to paleopathology has mainly focused on infectious diseases, such as confirming *Yersinia pestis* as the pathogenic agent of ancient plagues (23). Comparing the genes related to pathogenic resistance in ancient populations (24). Determining the oral and microbiome of ancient individuals (25). Or studying the prevalence of malaria in ancient times (26). However, Ancient DNA has a huge potential to understand the genetic architecture of diseases affecting ancient populations (27). With this idea in mind, I used genomic data for 80 ancient individuals for which we know who were affected or not by PH as well as hundreds of SNPs related to specific phenotypes to test for the different hypothesis proposed for the origin of PH (see Table 1 for the hypothesis tested and its predictions).

**Table 1.**
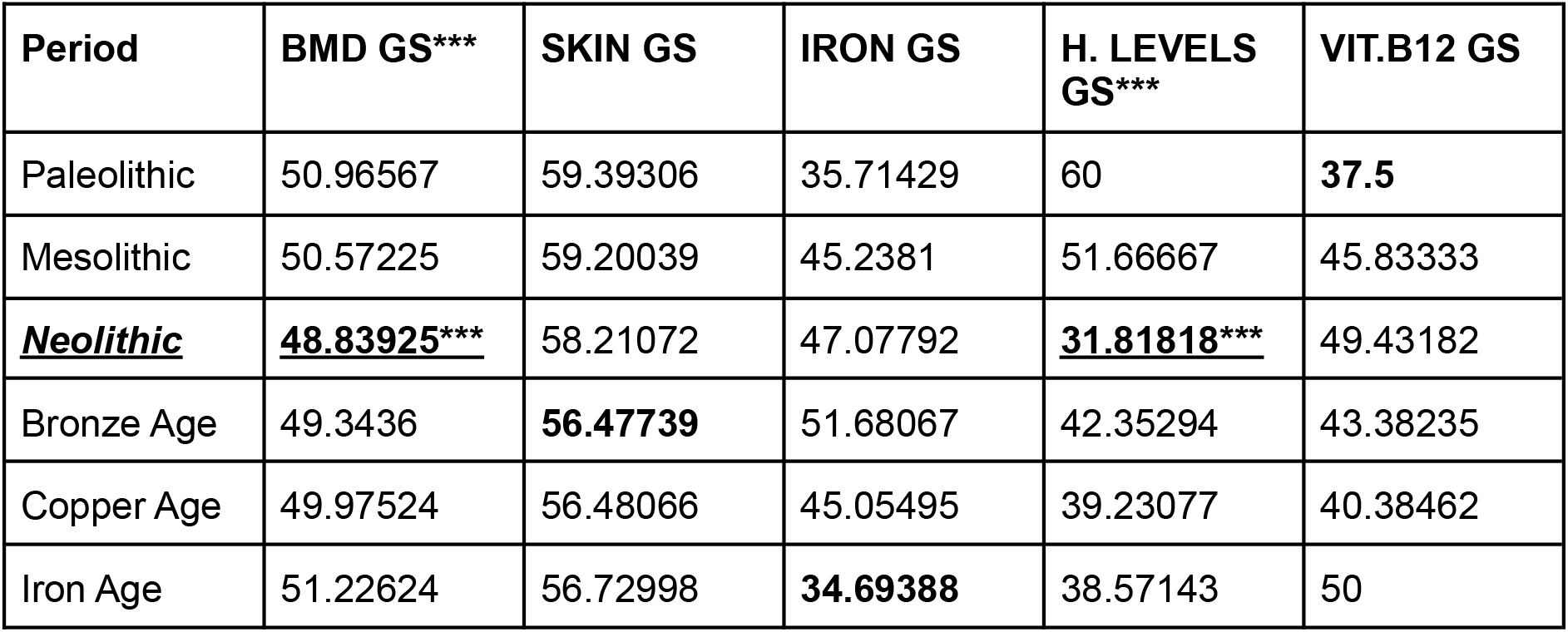
Genetic risk score for the different phenotypes tested here: Bone mineral density, Skin pigmentation, Iron metabolism, Haemoglobin levels and Vitamin B12 metabolism. In black, the lowest value in each phenotype. *** marks the phenotypes where the lowest values coincide with the Neolithic (Bone mineral density & Haemoglobin Levels).

| Period | BMD GS*** | SKIN GS | IRON GS | H. LEVELS GS*** | VIT.B12 GS |
| --- | --- | --- | --- | --- | --- |
| Paleolithic | 50.96567 | 59.39306 | 35.71429 | 60 | **37.5** |
| Mesolithic | 50.57225 | 59.20039 | 45.2381 | 51.66667 | 45.83333 |
| **<u>Neolithic</u>** | **<u>48.83925\*\*\*</u>** | 58.21072 | 47.07792 | **<u>31.81818\*\*\*</u>** | 49.43182 |
| Bronze Age | 49.3436 | **56.47739** | 51.68067 | 42.35294 | 43.38235 |
| Copper Age | 49.97524 | 56.48066 | 45.05495 | 39.23077 | 40.38462 |
| Iron Age | 51.22624 | 56.72998 | **34.69388** | 38.57143 | 50 |

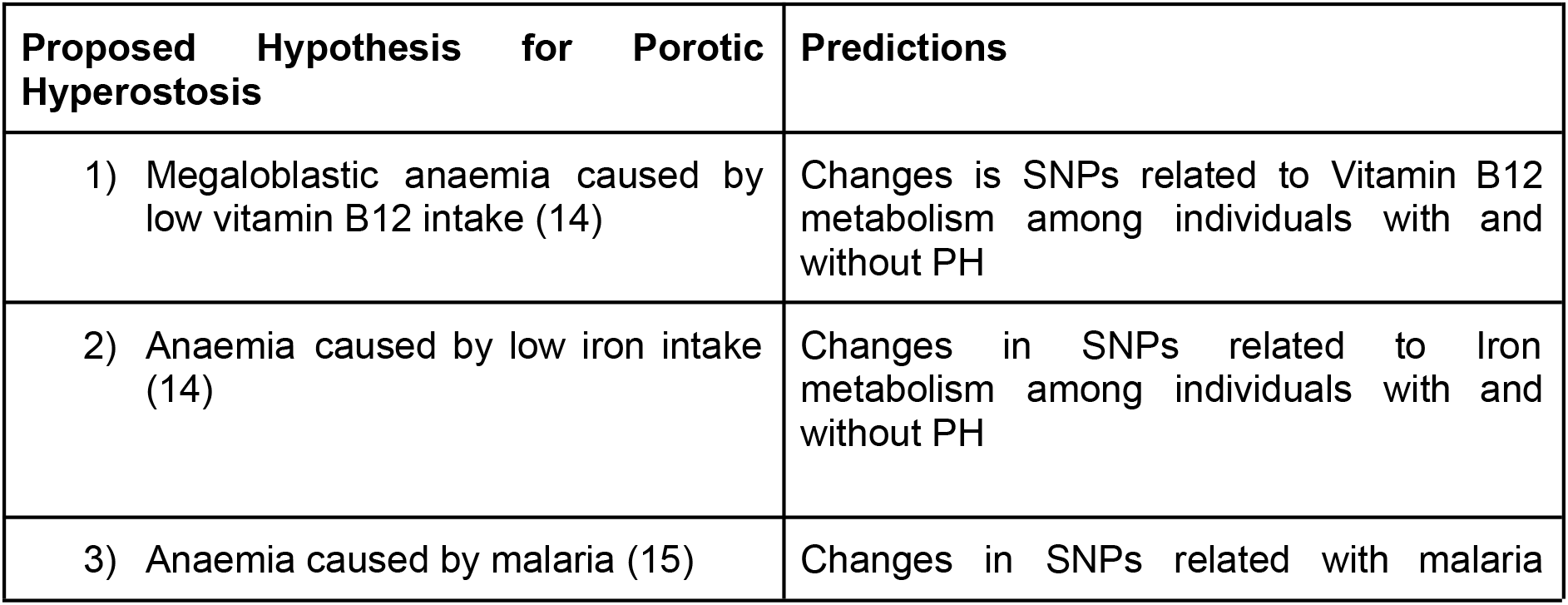
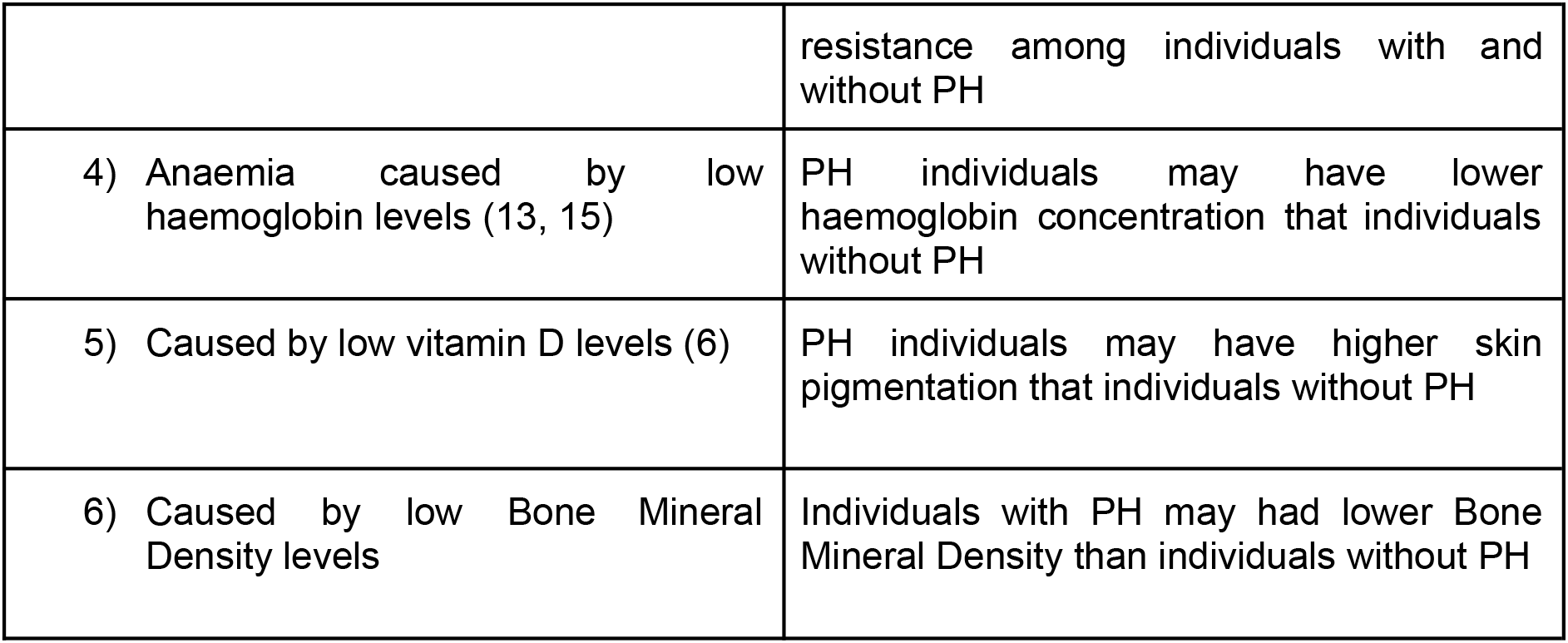

The results suggest anaemia as the main cause for PH. Specifically, an anaemia that was caused by a genetic tendency for low haemoglobin levels together with genetic architecture for low bone mineral density. Specifically, Neolithic individuals show low levels for BMD compared to previous and recent populations which may explain why PH had the highest prevalence levels during this period.

## 2. Material and methods

### Databases

#### SNPs related to BMD

Bone mineral density is the amount of minerals (such as calcium) deposited in the bones. Many factors influence the levels of BMD. It can be influenced by environmental cues like the levels of diary activity or the diet, but also by genetic factors (16, 17). Eurasians tend to have lower BMD compared to African populations. This has been interpreted as the result of selective events since the Out-of-Africa (16, 17). In order to see if individuals with PH had lower BMD I used the most broad study of SNPs related to BMD, with more than 1000 SNPs (28). The datasets used here recover 233 of these SNPs (see Supplementary Table 1). I used these SNPs to create a genetic score of the BMD in the ancient individuals.

#### SNPs related to skin pigmentation

Europeans evolved low levels of melanin in the skin cells. However, ancient DNA studies suggest that this selection increased after the Agricultural Revolution (29, 30). Despite the main evolutionary reason remains unknown, low melanin production has been related to increased levels of vitamin D in blood. Vitamin D is synthesised by the influence of UV-light from the sun and skins with higher melanin production blocks most of this UV-light (30). Vitamin D is needed to absorb calcium in the guts and is then fundamental to develop healthy immune and skeletal systems, including normal levels of mineral deposition in the bones (30).

As selective pressure for light skin increased during the Neolithic (29, 30), early farmers may have had lower vitamin D levels than recent populations (such as Bronze, Copper or Iron age individuals) and then a lower capacity to synthesise vitamin D. This may explain why the prevalence of PH was higher during the Neolithic than during recent times. To test for it I used a broad dataset of SNPs related to skin pigmentation (that has previously applied to ancient DNA studies (30)) to generate a genetic score of skin pigmentation in the ancient samples. 173 from 175 SNPs related to skin pigmentation were recovered in the dataset used here (see Supplementary Table 2).

#### SNPs related to iron metabolism

Iron is needed to produce haemoglobin, the protein that transports oxygen molecules throughout our body. Its deficiency usually leads to anaemia and may be the cause of the expansion of the cranial diploë. Several PH studies suggest that a diet low in iron may have followed the transition from hunter-gathering to farming. Also pathogens can retain the iron from our blood reducing its levels. It has also suggested an increase of pathogenic diseases during the early Neolithic, then reducing the levels of iron in early neolithic individuals. This may explain why PH had a higher incidence than in recent times. All these scenarios should have triggered changes in the genes related to iron metabolism. To test for this idea I study changes in the genetic score values in 8 SNPs (31) related to iron levels in Europeans that can be recovered in the ancient dataset used here (see Supplementary table 3).

#### SNPs related to vitamin B12 metabolism

Vitamin B12 is needed for a normal formation of red cells. Animal diet is richer in Vitamin B12 than a diet rich in vegetables. Transition from hunter-gathering to farming may then have led to a diet low in vitamin B12 and an impaired formation of red cells. This can explain why PH had a higher prevalence than in previous times (Mesolithic and Palaeolothic) and also in recent times as animals were domesticated and included in the diet. This situation should have triggered changes in vitamin B12 metabolism. Here I used a genetic score with 4 SNPs (32) related to vitamin B12 metabolism that can be recovered in the database of ancient samples used here (see Supplementary Table 4).

#### SNPs related to malaria resistance

It is still controversial how prevalent malaria was in ancient populations (26, 33). *Plasmodium falciparum* is a parasite that destroys red cells during its life cycle, leading to anaemia. Several studies point toward malaria as the cause for PH. To test for this hypothesis I used 104 SNPs related to malaria resistance (34), together with 22 SNPs from (26). As this phenotype can be caused by a single genomic variant, I decide not to conduct a genetic risk score in this phenotype but a FST analysis as in Gelabert. P. et al. 2017 (26), where the authors used 22 SNPs to look for evidence of malaria as a selective agent in ancient populations. 77 of all these SNPs could be recovered in the ancient datasets (see Supplementary Table 5).

#### SNPs related to haemoglobin levels

Haemoglobin is the protein that binds oxygen molecules inside the red cells. Low levels of haemoglobin lead to anaemia and then may cause PH. Low levels have been related to genetic variants (35) such as thalassemia, a genetic condition affecting one of the genes for the α of β globin (35). Interestingly, thalassemia is common around the Mediterranean sea populations and has been referred to as Mediterranean anaemia. To test for low haemoglobin levels as the main cause for PH, I conducted a genetic score with SNPs related to haemoglobin levels in inhabitants of the Sardinian island (west part of the Mediterranean sea) (36). 5 SNPs could be recovered in the ancient dataset, one of them affecting the gene for the β globin (see Supplementary Table 6).

### Ancient datasets

For the main analysis I took data about 47 individuals for which it is known that they didn’t have PH and 33 individuals that had PH (see Supplementary Table 7). These 80 individuals belonged to a dataset of 167 ancient individuals (37) (see Supplementary table 8). The whole dataset was later used together with another dataset of 153 ancient individuals to see changes in the BMD and Haemoglobin levels in Europeans during the last 40,000 years (38) (ee Supplementary Table 9).

### Genetic risk score estimation

Many phenotypes are influenced by multiple genetic variants, as for example bone mineral density or skin pigmentation. All the additive effects of each variant can be aggregated together into a simple measure, known as genetic risk score (39). Genetic risk score can be understood as a value for the genetic risk of an individual to develop a particular phenotype. It has been continuously applied to ancient individuals, for example the two datasets used here independently used a genetic risk score to calculate the height of the individuals based on height related SNPs (37, 38). Other studies applied genetic risk scores to measure skin pigmentation as in Du, J. & Mathieson, I. 2020 (30) or the likelihood to develop attention/deficit hyperactivity disorder in Neanderthals and ancient Europeans (40). As well as other hundreds of different phenotypes (41). The genetic risk score here was calculated as the percentage in each individual of SNPs related to: 1) high Bone Mineral Density, 2) darker skin pigmentation, 3) high iron levels, 4) high vitamin B12 levels and 4) high Haemoglobin levels (see Supplementary Table 10 & Supplementary Table 11).

### FST analysis for malaria resistance

FST analysis for the 77 malaria resistance SNPs were calculated with PLINK (42) (see Supplementary Table 5).

### Statistical analysis

All statistical analyses were conducted in R Studio (43). Wilcoxon Mann Whitney test was used to analyse differences in individuals with and without porotic hyperostosis among the different genetic scores. Pearson correlations were used to study differences among the genetic scores of BMD and Haemoglobin levels through time.

### Figures and plots

Figure 1 was generated in Biorender (44), other figures were conducted in RStudio (43).

## 3. Results and discussion

### 3.1. Porotic hyperostosis is common in ancient individuals with a genetic risk for low bone mineral density and low haemoglobin levels

When comparing the individuals with and without PH I detect statistical differences for the genetic risk scores of BMD (see Figure 2) and Haemoglobin levels (see Figure 3) (the p values for Mann Whitney Wilcoxon tests were 0.00007605 for the BMD and 0.02489 for the Haemoglobin levels). Contrary, the genetic risk scores of skin pigmentation, iron levels, vitamin B12 levels did not show statistical differences among the two groups. Individuals with PH had lower genetic risk scores for a high BMD during all ages from the Neolithic to the Copper Age (see Figure 2). On the other hand, the genetic risk scores of Haemoglobin levels show statistical differences only when all individuals are compared and during the Neolithic, but lose significance during the Bronze and the Copper Ages (Figure 3), this may be due to the lack of data as only 5 related SNPs could be used in this analyses. Palaeolithic individuals were not used in this analysis due to the low number of samples. Iron Age samples were also not used as no individual has PH.

**Figure 2.**
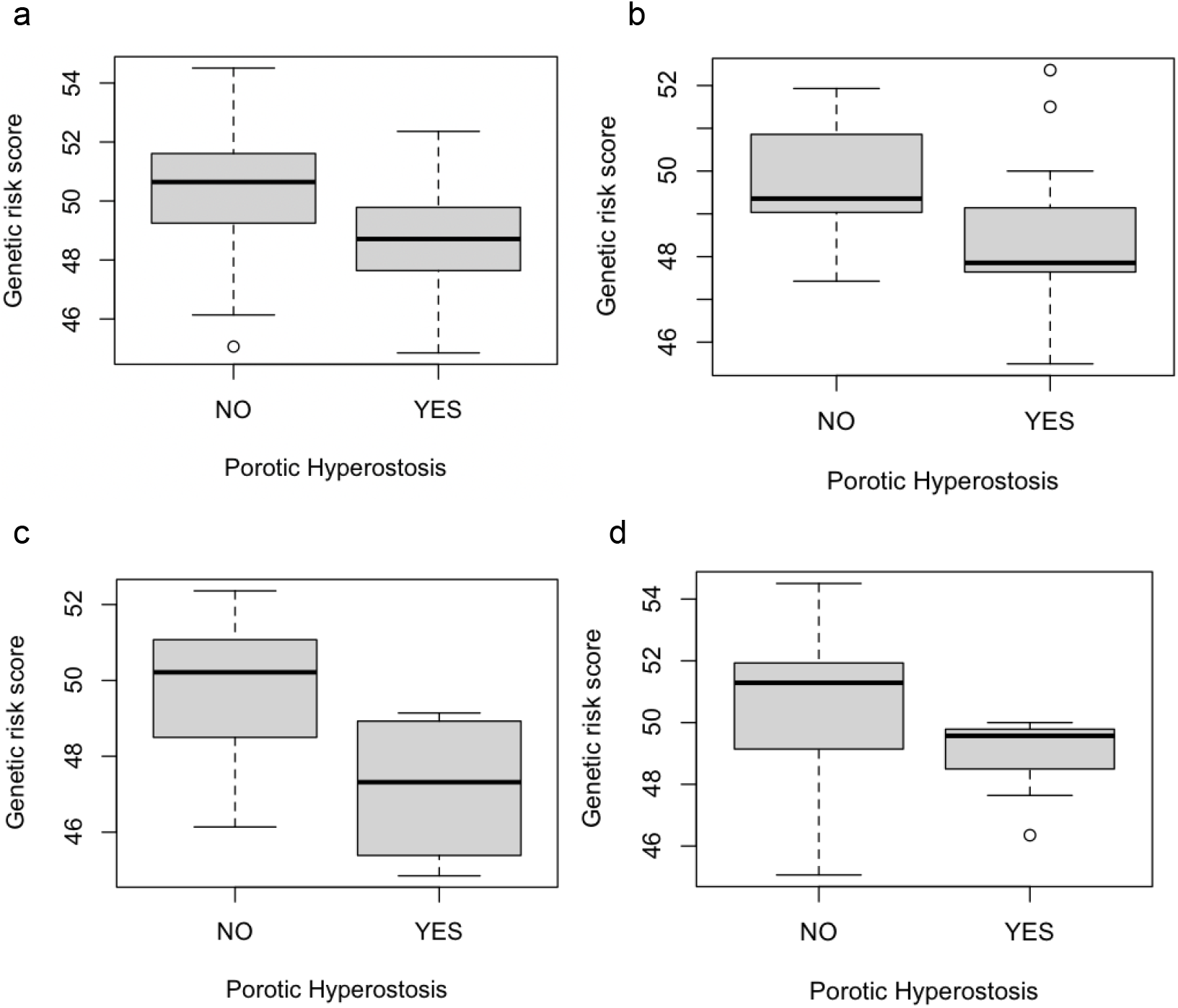
Individuals with porotic hyperostosis tend to have lower frequency of high BMD related alleles in all observed ages. a) Whole dataset, p value = 0.00007605. b) Neolithic p value = 0.1041. c) Bronze age, p value= 0.05366. d) Copper age, p value = 0.03521.

**Figure 3.**
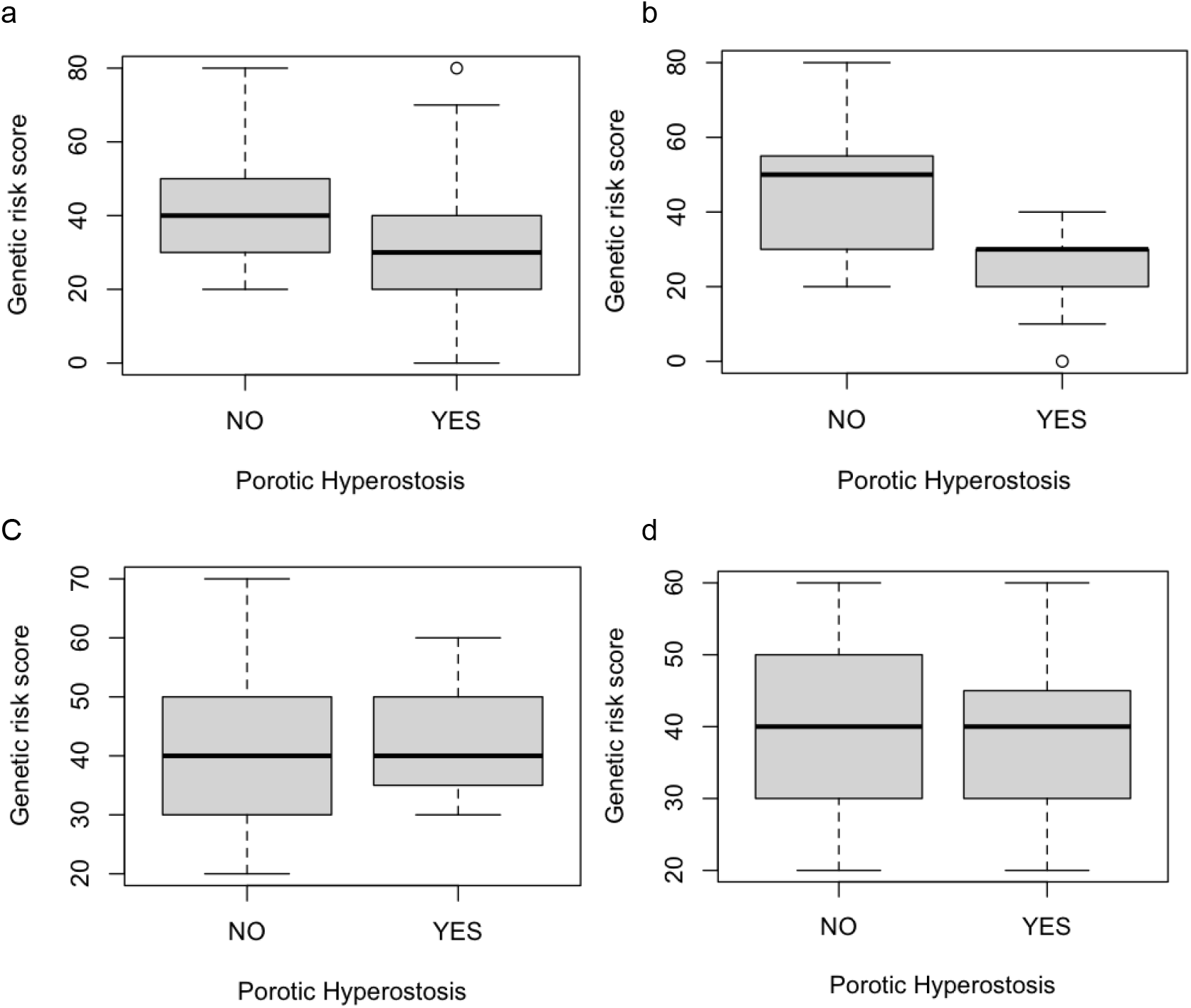
Individuals with porotic hyperostosis tend to have lower genetic risk scores for haemoglobin levels, specifically during the Neolithic. a) Whole dataset, p value = 0.02489. b) Neolithic p value = 0.03555. c) Bronze age, p value= 1. d) Copper age, p value = 0.8111.

**Figure 3.**
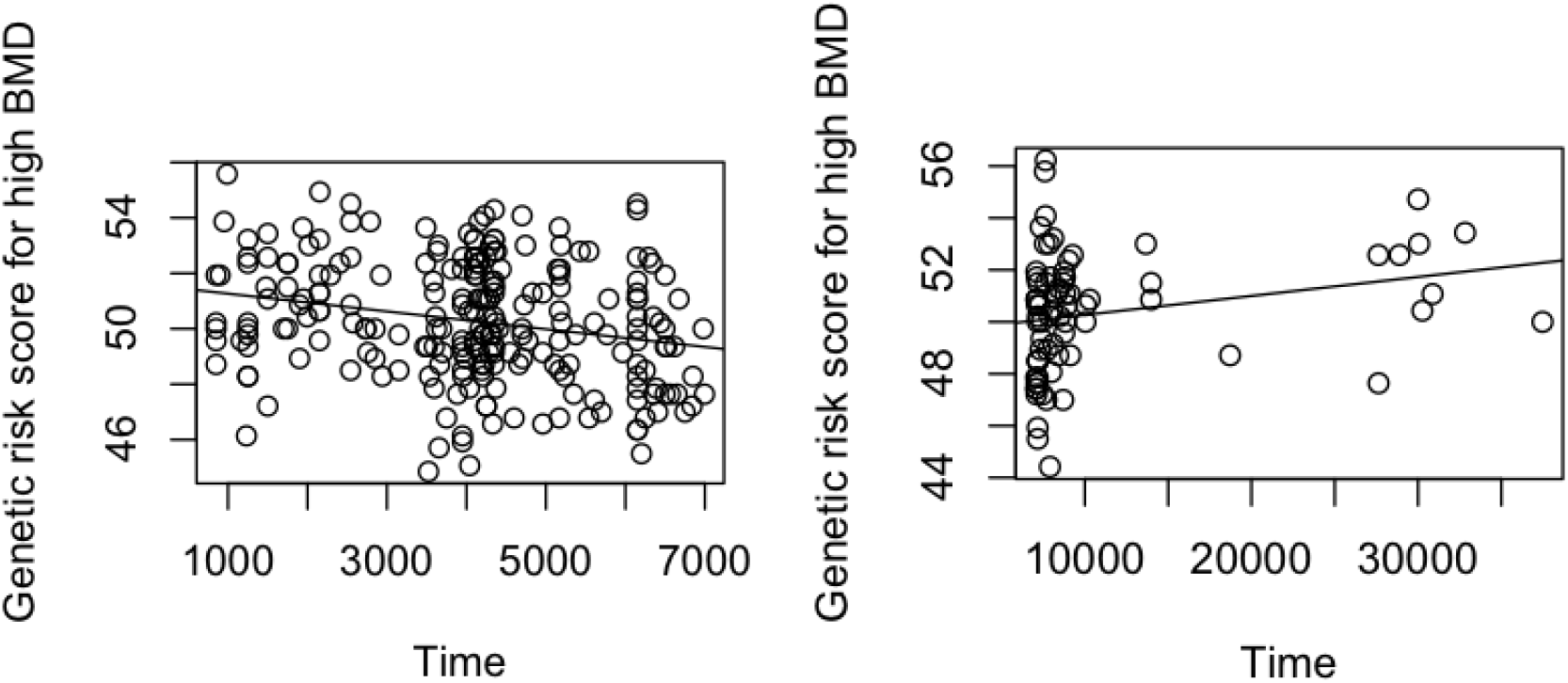
a) A statistical increment in the BMD risk score has been observed since the Early Neolithic to the Iron age (p value= 0.0002878, R2 = 0.05568). Only individuals younger than 7033.357 years ago are plotted. b) A statistical decay of the BMD risk score has been observed since the Paleolithic to the Neolithic (p value = 0.04012, R2 = 0.05645). Only individuals older than 7033.357 years ago are plotted. Each dot represents an individual.

These results do not support the hypothesis that PH was caused by low vitamin D levels in darker individuals. Neither changes in the iron or in the vitamin B12 metabolism. Instead, support the hypothesis that anaemia caused by low haemoglobin levels may be responsible for the PH, together with a genetic architecture for low bone mineral density. Interestingly, low BMD correlates with low haemoglobin levels (45). A possible explanation for this is that anaemia caused by low haemoglobin levels may have correlation with a genetically porous bone to allow for an expansion of the diplöe.

### 3.2. Neolithic individuals had the lowest values for Bone mineral density and low Haemoglobin levels

In order to understand what could have caused such prevalence of PH in the Neolithic, I calculate the mean of the genetic risk scores by time periods (from the Palaeolothic to the iron Age) (Table 1). The genetic scores for skin pigmentation and iron and vitamin B12 metabolism had lowest mean values in other periods but in the Neolithic. However, both genetic risk scores for BMD and for haemoglobin levels show the lowest mean values during the Neolithic. In particular the genetic risk score for BMD has decreased since the Palaelothic to the Neolithic, and increased again from the Neolithic to the Iron Age.

This last result is confirmed by studying the genetic risk scores of 308 (including the 80 used in the previous analysis) ancient samples (see Materials & Methods) through time (Figure 4). The mean value for the age of the Neolithic individuals with PH was 7033.357 years. In order to confirm that BMD decreased from the Palaeolothic to the Neolithic I compared the genetic risk score for the BMD levels of all the individuals older than 7033.357 years ago. I saw a statistical decreasement in the genetic risk score for the BMD reaching the lowest values in the Neolithic period (p value = p value = 0.04006, R2 = 0.05725). On the other hand, I test for an increment of the genetic risk score of the BMD after the Neolithic by comparing all the individuals younger than 7033.357 years ago. The results show a statically increment of the genetic risk score for the BMD with the lowest values being observed in the Neolithic (p value= 0.0002707, R2 = 0.05592). All these results suggest that Neolithic individuals had the lowest risk scores for BMD and can explain why PH had this high prevalence in this specific period compared with previous and later times (2, 9-11).

**Figure 4.**
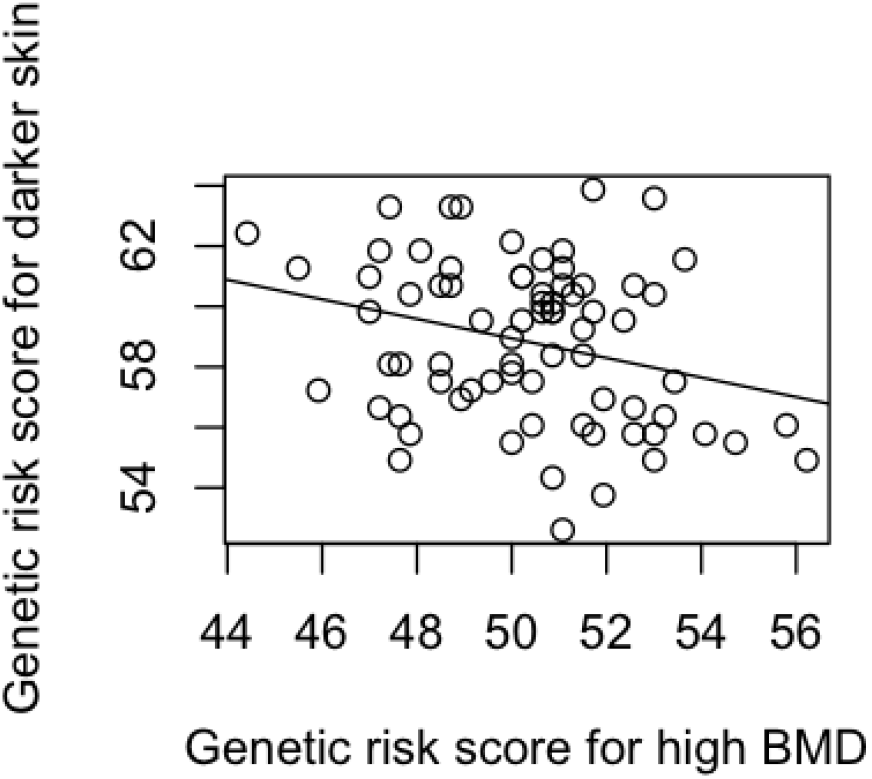
The genetic score for darker skin pigmentation correlates with the genetic score for high BMD levels in 74 ancient individuals belonging to the Early Neolithic or previous periods. In the x the genetic risk score for the BMD, in the y the genetic risk score for darker skin pigmentation. Each dot represents an individual.

The observed decrement in the BMD during the Palaeolithic and Mesolithic is compatible with previous studies suggesting a selection for a low BMD in Eurasians during the last thousand of years (16,17). As selection for light skin increased drastically after the Neolithic in Europeans, low BMD levels may be an adaptation to low UV-radiation in the European latitudes. In this line I observed a statistical correlation among the genetic risk scores for skin pigmentation and BMD in ancient mesolithic and palaeolithic individuals (Figure 4) (p value = 0.01368, R2 = 0.0815), where low genetic score for BMD correlates with high genetic scores for a darker skin pigmentation. However, this correlation is not observed after the Neolithic (p value = 0.6766, R2 = 0.0007544).

## 4. Conclusion

Porotic hyperostosis was a common disease in ancient times, specifically during the Neolithic. It is commonly accepted that anaemia is the main cause for this disease, however, genetic analysis has not been tested in ancient remains. Here I tested several proposed hypotheses using 80 ancient individuals for which we know if they had or not PH, belonging to different periods from the Palaelothic to the Iron age. Despite environmental factors can influence the development of PH, the results suggest that the individuals may already be genetically predisposed for the disease. I do not observe statistical changes in iron or vitamin B12 metabolisms, neither in malaria resistant alleles or in the vitamin D levels. On the other hand, I confirm that individuals with PH had a genetic disposition to low haemoglobin levels as well as for a low bone mineral density. Both phenotypes are known to correlate, which is likely due to the fact that red cells generate in the diplöe, which grows inside the bone matrix. Interestingly, the low BMD observed in the Neolithic compared to all other periods can explain the highest prevalence of the PH during the Neolithic. BMD seems to have decreased from the Palaeolothic to the Neolithic and increased again after the Neolithic. A possible explanation for the decrease is that ancient Europeans adapted to the low UV-radiation by reducing the need for calcium for the bones (reducing the BMD), however I am unable to explain how the BMD increased again after the Neolithic.

## Data Availability

All data produced in the present study are available upon reasonable request to the author

## Acknowledgements

I would really like to thank Dr. Ian Mathieson for his kindness in giving me access to the dataset of the 154 ancient individuals.

## Notes

### Competing Interest Statement

The authors have declared no competing interest.

### Funding Statement

This study did not recieve any funding

### Author Declarations

The study used genomic data from ancient individuals that has been already published in other two studies.

